# Virtual wards: A rapid evidence synthesis and implications for the care of older people

**DOI:** 10.1101/2022.06.24.22276864

**Authors:** Gill Norman, Paula Bennett, Emma R.L.C. Vardy

## Abstract

Virtual wards are an area of rapid development within the National Health Service in the UK, and frailty has been selected as one of the first clinical pathways to be developed. This is in the context of existing longstanding hospital at home services in some areas. A rapid evidence synthesis was completed to guide the local healthcare system in the North West of England. This is a rapid approach to synthesising existing research which focuses on existing evidence synthesis where possible. Questions were addressed on clinical-effectivness, cost-effectiveness, barriers and facilitators, models of care and use in practice, and staff and patient and carer experience. We found that whilst there was a significant evidence base for hospital at home, there was less evidence for virtual wards. There is lack of guidance for key aspects of virtual wards including team characteristics, outcome selection and data protection. We recommend that research and evaluation is integrated into development of virtual ward models of care. Of particular relevance for older people cared for on virtual wards is the issue of carer strain, an area which may be frequently overlooked.

## Introduction

The concept of virtual wards has existed for some time. During the pandemic the use of virtual wards was expanded with apparent good effect to manage selected patients with COVID-19 using a pulse oximeter and monitoring through secondary care.(2) Subsequently large scale investment is being made to support virtual ward expansion in the NHS to include patients with frailty.

To support integrated care systems and service providers to establish and expand virtual wards, two high priority pathways have been introduced,(3) acute respiratory tract infection virtual wards and hospital at home for those with frailty.(4) This is a seismic shift to the way healthcare is to be delivered for older people, and an area which is currently being prioritised for funding.

It is important to clarify terminology at this stage given the existence of hospital at home and other community led services alongside virtual wards; this is summarised in Box 1.

### Virtual wards, hospital at home and remote monitoring

- **Hospital at home** services provide face-to-face care at home through a multidisciplinary team (MDT) based in the community. They are provided as an alternative to inpatient care.(1)
- **Virtual wards** are a hospital-led alternative to in-patient hospital care and may incorporate remote monitoring through apps, technology platforms, wearables and devices such as pulse oximeters.(2) In their simplest form remote monitoring represents most or all of the provision in virtual wards; this was exemplified by provision of pulse oximetry to Covid-19 patients.(5)
- **Remote monitoring** includes virtual wards but is a broader term and is not always restricted to people who would otherwise require inpatient hospital care.
- **In practice** there is often considerable overlap between hospital at home and virtual wards; remote monitoring is often used in conjunction with care provision; the terminology sometimes used “virtual wards (hospital at home)” reflects this. (7)
- **There is a continuum of care provision** in conjunction with remote monitoring in virtual wards and this overlap is greatest where care needs are highest – in older, frailer patients with long-term conditions. (6)

**Box 1: Services and terminology used in practice**

The British Geriatrics Society has previously published guidance outlining many of the principles for successful home based services.(6) In order to inform wider implementation of virtual wards in Greater Manchester in the North West of England we carried out a rapid evidence synthesis (RES) of existing systematic reviews of virtual wards, hospital at home and remote monitoring as alternatives to acute hospital admission or stay. Because of the close relationship between virtual wards and both hospital at home and remote monitoring we searched for, and included, systematic reviews relating to any one of these, where the service was provided as an alternative to inpatient hospital admission. We summarise and contextualise the findings here; for the full RES, including all references, please see supplementary information (Appendix 1).

### Objectives

to rapidly synthesise evidence from existing evidence syntheses which was relevant to the clinical and cost-effectiveness of virtual wards; the barriers and facilitators to their use; the ways in which they are used; and the experience of patients, carers, and staff.

## Methods and Search Strategy

We employed a methodology outlined in our RES framework, described briefly below. (8, 9) The following eligibility criteria were used:

1. **Population:** People who would otherwise require acute hospital inpatient care. People who required acute mental health care were excluded. We prespecified the following subgroups as being of particular interest: people with acute respiratory conditions including chronic obstructive pulmonary disease (COPD) exacerbations or COVID-19; people with heart failure; people with frailty, but did not restrict the RES to reviews focusing on these conditions.
2. **Intervention:** Hospital at home; virtual ward; remote monitoring. We included both step-up (hospital admission avoidance) and step-down (hospital early supported discharge) models.
3. **Comparator:** Acute inpatient care
4. **Outcomes:** Outcomes are reported for each key question below
5. **Study design:** Systematic reviews

We accepted authors’ definitions of the populations, interventions, comparators, and outcomes; we required reviews to have systematic searches and clear inclusion criteria.

In March 2022 MEDLINE OVID, CINAHL-PLUS EBSCO, and the Cochrane Database of Systematic Reviews were searched using a strategy devised by an information specialist based on the interventions of interest (supplementary material). We also searched medRixv for relevant preprints and checked the references of identified reviews.

## Result

The search identified 630 unique records. Following full-text screening of 52 records, 39 publications relating to 31 unique reviews were included.(1, 10-39) This included four Cochrane reviews.(1, 10-12) Although most included reviews did not use age or frailty as inclusion criteria, most participants included in the reviews were older and/or had one or more chronic conditions, and the identified Cochrane reviews were included in a rapid review of hospital at home as a component of the frailty pathway.(39) We therefore consider the results of the RES as a whole to be relevant to the use of virtual wards for older people.

### Clinical-effectiveness

We looked at the following outcomes: Length of stay in any setting (hospital or virtual ward), readmission, mobility, need for community support after discharge; achievement of rehab goals, adverse events including mortality, unplanned contacts/treatment events, acceptability (to patient/carer/staff), satisfaction (patient/carer/staff). We report acceptability and patient satisfaction together with experience below.

There was consistent moderate or low certainty evidence from Cochrane reviews that hospital at home probably results in most clinical outcomes, including mortality, being as good or better than inpatient care, for both step-up models of admission avoidance and step-down models of early discharge.(1, 10) We particularly noted that there was probably a reduced rate of admission to residential care following treatment at home in either step-up or step-down models.(1, 10) An exception was length of stay, which was longer in the step-up model,(1) although shorter in the step-down model.(10)

The review of step-up care included 16 RCTs with 1814 participants,(1) and the review of step-down care included 32 RCTs with 4746 participants.(10) The judgements that the evidence was low or moderate certainty represent the GRADE assessments of the Cochrane review authors and mean that the effect estimates probably are (moderate certainty) or may be (low certainty) close to the true effects, but that further research may change the effect estimates for the outcomes, and may change the direction of an effect.(40) These findings were supported by other Cochrane and non-Cochrane reviews in both general,(13-18) and condition-specific populations.(11, 22-24, 26-29) While there was substantial evidence for mortality, length of stay, and admission to residential care, we found more limited information on other outcomes. There were also some differences in the evidence for different patient groups and between step-up and step-down models; these are explored in more detail in the full RES (see supplementary information in Appendix 2). For example, people with COPD in particular may have reduced readmissions after hospital at home, (10, 11, 15, 22) while those with a mix of acute medical conditions may have an increased risk of readmission after step-down hospital at home.(10) Although there were three reviews of remote monitoring in COVID-19 the primary studies in these reviews did not have designs which enabled robust evaluation of effectiveness.(19-21)

### Cost-effectiveness

We looked for measures of cost-effectiveness (e.g. QALY) and relative cost-effectiveness (e.g. ICERs) but also reported cost measures. The evidence we identified relates mostly to hospital at home models.(33) Although most primary studies showed estimated cost-savings from hospital at home these estimates vary widely, ranging from savings per patient of over EUR 8000 to increased costs of over EUR 2000 indexed to 2018 prices.(33) Importantly, most studies used methodologies which meant that they were likely to overestimate cost-savings. Quality assessment showed an average score of 60 out of 100 points, with almost all failing to meet one or more criteria for avoiding risk of overestimating savings. In particular, many studies used a generic unit price for inpatient days, rather than reflecting disease or unit-specific costs or the decreasing care intensity (day specific costs) across a stay. Therefore, the cost-effectiveness of treatment at home is uncertain despite the large number of studies evaluating it.

Importantly studies disregarded costs to patients and carers. Despite 21 of 48 economic studies using availability of informal care as an inclusion criterion only two included costs of informal care in their analysis.(33) Studies which did consider costs to families included paid and unpaid domestic help and personal care, including the time of the informal caregiver.(33) One study estimated the mean per patient additional costs associated with informal care for COPD patients treated at home at over EUR 500 (2009 reference year) more than for those treated as inpatients in the Netherlands over a seven day treatment period and three month follow-up.(41) The disregarding of such costs in most studies may have implications for the applicability of research to disadvantaged groups. This should be taken into account when considering the Cochrane review finding that step-up models may be less expensive than inpatient admission, excluding informal care costs.(1)

### Barriers and facilitators

We considered all factors at both the patient/carer level and the staff/system level. We included barriers to and facilitators of setting up of virtual wards/hospital at home as well as to enrolment in them. We were particularly interested in patient-level factors with implications for equity such as digital literacy.

#### Organisational and interventional

Many of the barriers to remote monitoring related to organisational or team characteristics, including the lack of guidance on team characteristics, data governance and organisational oversight. Identification of the appropriate clinical outcomes for patient monitoring was also identified as a concern by staff. (38) Facilitators at the organisational level related to supportive operational, regulatory and legal frameworks, co-ordination and integration of care, including with post-discharge care; and staff with strong clinical and communication skills. (15, 34, 38)

Interventions which were tailored to patient conditions and situations were associated with successful implementation. Having patient involvement in design of remote monitoring interventions was identified as a factor in their success. Making the intervention simple and easy to use; ensuring accurate and sensitive measurements; using patient-specific measurements; and co-ordinating the intervention with self-management (e.g. monitoring of medication adherence), combined with support, education, and feedback to personalise care were also considered important to success.(38)

#### Interpersonal and Intrapersonal

Many of the facilitators identified related to patient characteristics. Patients were selected for factors such as strong social support, positive health behaviours, confidence in receiving care at home, and conducive home environment.(34) Patient and carer training was also associated with successful remote monitoring.(19) Barriers to implementation included greater physical distance of patients’ homes to the hospital, medical condition stability and level of disability.(15) Issues around health and technological literacy and access to internet or internet-enabled devices were identified as barriers; use of telephone-based monitoring was considered to be more inclusive for some patients. Equity-related factors were the largest group of barriers in one review, the most commonly identified of these was place of residence, including rural or remote residence, nursing home residence or homelessness.(21) The fact that patient characteristics were so strongly identified as both barriers and facilitators may indicate that careful consideration of patients’ characteristics before enrolment is important to successful use of virtual wards. It also suggests that there may be limits to the applicability of virtual wards to some groups of patients.

### Models of care and use

This question looked at how hospital at home and virtual wards are used, including whether step-up/step-down models are used, and the characteristics of people admitted to them. As with barriers and facilitators we paid particular attention to equity considerations. We also looked for evidence comparing the effectiveness and cost-effectiveness of different models of care including the use of different levels of patient engagement or continuous versus intermittent monitoring.

Both step-up and step-down models of hospital at home were assessed in separate Cochrane reviews.(1, 10) Step-up models of care treated patients referred from emergency departments, outpatients and primary care.(1) While one overview of reviews looked at these two models together (15) we did not identify other comparisons of different models of treatment at home: there were no comparisons of virtual wards with traditional hospital at home, or with different approaches to virtual ward delivery. The level of multidisciplinary input in teams providing care varied; where non-acute care needs were higher there was more nurse-led care and more family involvement in care; higher levels of acute care need were associated with involvement of a greater range of healthcare professionals.(35)

Reviews and the primary studies they included often had restrictive, sometimes condition-specific inclusion criteria for participants. Most people in the primary studies were older (in the Cochrane review of step-up hospital-at-home mean participant ages ranged from 70 to over 80 years(1)) and/or had one or more chronic conditions; in some cases the treatment need resulted from an exacerbation of this condition, in others an additional acute condition was the proximal cause of admission.

Specific conditions which we identified reviews in were: COVID-19, (19-21) COPD,(11, 22, 23) heart failure,(26-29) pulmonary embolism,(24) and people at the end of life. (12) People recovering from stroke and people recovering from surgery were also represented as prespecified subgroups in Cochrane reviews while many studies enrolled people with a mix of acute medical conditions.

In the Cochrane review of step-up care one trial included only frail older people with dementia. (1) However many of the participants across the reviews would be likely to be classed as frail although they were not specifically included on that basis As we noted, a review which considered hospital at home as a component of the frailty pathway used the Cochrane reviews we identified (1, 10) as their main evidence source.(39)

### Staff and patient experience

We looked at all measures of patient or carer experience, satisfaction, and acceptability. We also report staff experience.

In mixed methods and qualitative studies expressed satisfaction with hospital at home is generally high amongst both patients and staff involved in delivery.(34) Patient satisfaction in reviews of randomised controlled trials (RCT) may be slightly higher in those treated in hospital at home compared to those treated as inpatients, although some studies find no difference.(1, 10) A recent review of qualitative studies found that the decision to have care at home was often determined by the preferences of healthcare professionals and patients, with less consideration given to the views of carers or families. Caregiver burden was, however, a theme in many of the identified studies, particularly where patients had dementia or mental illness; some caregivers were described as having “burnt out” during hospital at home.(34) Carer outcomes in intervention studies may not reflect these perspectives, with some reporting reduced stress.(10) Advantages, disadvantages and challenges identified in the perspectives of both staff and patients (34) reflected elements identified as influencing success of remote monitoring interventions,(38) but the need to consider the experience of informal carers was highlighted in this meta-synthesis.

## Discussion

### Strengths and limitations of this work

This systematic search identified a substantive body of existing evidence syntheses relevant to the area. However, the methods used are not those of a full systematic review, they are an adaptation of methods designed to provide a very rapid summary of the evidence to support local decision-making and planning around innovation implementation, and to inform further evaluation. Primary research is likely to exist which would fill in gaps, or complement the evidence identified here. Many of the included reviews are recent, nevertheless primary research published after the search dates of the reviews is not represented in this synthesis.

There is a more developed evidence base, especially in terms of clinical effectiveness, for hospital at home than for virtual wards; because the service model has been in use for longer there are many more trials and more systematic reviews. The evidence for the broader area of remote monitoring is also wider than virtual ward use. Systematic reviews of virtual ward use in COVID-19 exist and are included here, but the primary studies are predominantly non-comparative designs, limiting their ability to address questions of clinical effectiveness.

The representativeness of the populations included in trials or other primary research studies varies, meaning that systematic reviews may not be fully reflective of people treated in virtual wards in routine practice; the RES may therefore not fully reflect clinical practice, particularly at a time of rapid service evolution.

### Implications for research and practice

This rapid synthesis identified that whilst there is a substantive evidence base for hospital at home there is a need for robust evaluation of virtual ward models of care, given the rapid expansion of their use following COVID-19. Existing reviews of virtual wards in COVID-19 have not identified primary studies with robust designs for assessment of effectiveness.(19-21)

There is lack of guidance for key aspects of virtual ward provision – including team characteristics, outcome selection and data protection. This suggests that development and dissemination of evidence-based guidance for service delivery is a priority. With regards to frailty there will be significant implications for workforce design to support hospital at home service development, and how technology might be helpfully utilised. Distinctions might also helpfully be made in service modelling and data collation between prevention of deterioration and management of long-term conditions, for example in the context of heart failure, versus management of acute deterioration.

The role of family and other informal carers is likely key to the successful implementation, due to services being frequently used by people who have existing care needs prior to acute illness. Carer burden and the risk of carer burnout were identified as key considerations but there is little evaluation of the impact, including the financial impact, on carers. Evaluations of virtual wards should include carer outcomes and experiences as a priority. Evaluations should also include rigorous assessments of cost-effectiveness as well as clinical effectiveness, and these should include direct and indirect costs to patients, carers, and families. Existing evidence suggests that clinical outcomes for hospital at home are largely comparable to those of inpatient care. The evaluation of virtual wards that share many features with hospital at home may therefore need to prioritise the other questions addressed here, of cost-effectiveness, barriers to implementation, and patient and carer experiences.

Research in this area is evolving rapidly with rigorous primary studies becoming available;(42, 43) ensuring that implementation of services is informed by this is a priority. Those developing and implementing virtual ward services should be aware of the research on barriers and facilitators and should consider both organisational/interventional and patient-level factors. Approaches such as living systematic reviews (reviews which are continuously updated) may prove useful in ensuring that practice is informed by a rapidly developing evidence base.

### Concluding remarks

Development of virtual wards and hospital at home is rapidly evolving. This rapid synthesis summarises some of the key considerations in service development. It also highlights where evidence is lacking and the importance of building robust evaluation into new models. There is a substantial evaluation opportunity given implementation at scale. Rapid evaluations, potentially using routine data, are likely to be informative in initial assessments of the impact of these changes in service provision. The importance of co-production and co-design with service users is emphasised as well as impact, financial and otherwise, on unpaid carers. With regards to frail older people, specific attention should be paid to inclusivity of services for people with dementia so that older people are not disadvantaged, in terms of either the quality of care or through digital exclusion.

## Data Availability

All data produced in the present study are available upon reasonable request to the authors. The manuscript contains a report of a rapid synthesis of previously published data.

## Acknowledgements

The authors are grateful to Sophie Bishop for designing and implementing the search strategy, to Chunhu Shi for assistance in identifying relevant syntheses and for commenting on this manuscript, to Jo Dumville and Peter Bower for assistance in drafting and editing this manuscript, and to Bradley Quinn for assistance in developing the questions addressed.

## Author contributions

EV conceived the idea for the manuscript. GN and EV wrote the first draft of the manuscript, which is based on a rapid evidence synthesis (RES) conducted by GN. PB had the idea for the RES, helped to develop the questions addressed in it with GN, and commented on and edited the manuscript.

