## Appendix 1 for "Virtual wards: A rapid evidence synthesis and implications for the care of older people"

**Supplementary material: search strategy for RES**

**MEDLINE**

Ovid MEDLINE(R) and Epub Ahead of Print, In-Process, In-Data-Review & Other Non-Indexed Citations, Daily and Versions 1946 to March 15, 2022

\#	Searches

1	(virtual adj3 ward*).ti,ab.

2	(remote* adj3 monitor*).ti,ab.

3	(virtual adj3 monitor*).ti,ab.

4	(home* adj3 monitor*).ti,ab.

5	hospital at home.ti,ab.

6	1 or 2 or 3 or 4 or 5

7	exp Meta-Analysis as Topic/

8	meta analy$.tw.

9	metaanaly$.tw.

10	meta-analysis/

11	(systematic adj (review$1 or overview$1)).tw.

12	exp Review Literature as Topic/

13	7 or 8 or 9 or 10 or 11 or 12

14	cochrane.ab.

15	embase.ab.

16	(psychlit or psyclit).ab.

17	(psychinfo or psycinfo).ab.

18	(cinahl or cinhal).ab.

19	science citation index.ab.

20	bids.ab.

21	cancerlit.ab.

22	14 or 15 or 16 or 17 or 18 or 19 or 20 or 21

23	reference list$.ab.

24	bibliograph$.ab.

25	hand-search$.ab.

26	relevant journals.ab.

27	manual search$.ab.

28	23 or 24 or 25 or 26 or 27

29	selection criteria.ab.

30	data extraction.ab.

31	29 or 30

32	Review/

33	31 and 32

34	comment/

35	letter/

36	editorial/

37	animal/

38	human/

39	37 not (37 and 38)

40	34 or 35 or 36 or 39

41	13 or 22 or 28 or 33

42	41 not 40

43	6 and 42

**CINAHL**

CINAHLPlus Date Run 17/03/2022

| S19 | S6 AND S18 |
| --- | --- |
| S18 | S12 NOT S17 |
| S17 | S13 OR S14 OR S15 OR S16 |
| S16 | (MH "Animals") |
| S15 | PT editorial |
| S14 | PT letter |
| S13 | PT commentary |
| S12 | S7 OR S8 OR S9 OR S10 OR S11 |
| S11 | TI ( systematic review or systematic overview ) OR AB ( systematic review or systematic overview ) |
| S10 | (MH "Literature Review+") |
| S9 | TI Metaanalys* OR AB Metaanalys* |
| S8 | TI Meta analys* OR AB Meta analys* |
| S7 | (MH "Meta Analysis") |
| S6 | S1 OR S2 OR S3 OR S4 OR S5 |
| S5 | TI "hospital at home" OR AB "hospital at home" |
| S4 | TI (home* N3 monitor*) OR AB (home* N3 monitor*) |
| S3 | TI (virtual N3 monitor*) OR AB (virtual N3 monitor*) |
| S2 | TI (remote* N3 monitor*) OR AB (remote* N3 monitor*) |
| S1 | TI (virtual N3 ward*) OR AB (virtual N3 ward*) |

**Cochrane Database of Systematic Reviews**

Date Run:	17/03/2022

ID	Search	Hits

#1	(virtual near/3 ward*):ti,ab,kw

#2	(remote* near/3 monitor*):ti,ab,kw

#3	(virtual near/3 monitor*):ti,ab,kw

#4	(home* near/3 monitor*):ti,ab,kw

#5	"hospital at home":ti,ab,kw

#6	#1 or #2 or #3 or #4 or #5
