## Appendix 2 for "Virtual wards: A rapid evidence synthesis and implications for the care of older people"

NIHR | Applied Research Collaboration Greater Manchester

Health Innovation Manchester

### Rapid evidence synthesis: virtual wards (hospital at home) for acute admissions

#### Summary

There is **consistent low to moderate certainty evidence from reviews of randomised trials** that clinical outcomes, including mortality and readmission, for patients treated in hospital at home are probably as good or better than those treated as inpatients. The involvement of technology and different healthcare professionals in the models assessed varies, in some cases this is determined by the care needed. The **evidence on cost-effectiveness in unclear**; although there have been many studies of costs these nearly all show methodological issues which may mean they overestimate cost-savings. There is **insufficient evidence** on the cost implications for patients and carers. Barriers and facilitators exist at the **organisational, clinical and patient/carer levels** and there **was consistent evidence** on these. Identified patient-level barriers are **likely to have equity implications**. People treated in hospital at home (outside of Covid-19) are **mostly older and/or have one or more chronic condition**s; many are frail. In step-up models many are identified from emergency departments. There is an evidence base for COPD and heart-failure patients which aligns with the wider evidence. There is **low-certainty evidence from reviews of randomised trials** that patient satisfaction may be improved by hospital at home compared to inpatient care; there is less evidence around carer experience but the need for carer engagement is identified as a priority.

- ***Q1. Evidence for effectiveness***: There is consistent low to moderate certainty evidence from high quality systematic reviews of randomised controlled trials (RCTs) that use of hospital at home probably does not result in increases in mortality compared to usual (inpatient) care, either when used as a step-up model to avoid admission or a step-down model to shorten admissions. The evidence is less consistent that virtual wards for (re)admission to hospital; there is evidence that they may reduce this in people with chronic obstructive pulmonary disease (COPD), but the impact otherwise may depend on the type of model implemented and the patient population. The use of virtual wards probably decreases the likelihood of being admitted to residential care. The evidence on length of stay is more mixed; in some cases this is reduced and in some it does not differ from inpatient care.

- ***Q2. Evidence for cost-effectiveness*:** There is uncertainty around the cost-effectiveness of hospital at home. A 2019 review of 48 studies found a wide range of results, from savings of over €8000 to increases in cost of over €2000 per patient – although most studies showed cost savings. Almost all studies showed methodological problems meaning that they risked overestimating the cost savings. Cochrane and other reviews showed an uncertain impact of step-down models but a trend towards cost-saving in step-up models. Disregarding of direct and indirect costs to patient and carer may be an equity consideration in this evidence.

- **Q3. Barriers and facilitators:** There is consistent evidence that facilitators of virtual ward use are coordinated multidisciplinary teams involving staff with strong communication and clinical skills, and organisational structures which promote care delivery at home and continuity and integration of care over time, including with post-discharge care. Barriers include lack of information on patient data security measures, the need for robust outcomes testing, and the need to identify the most effective way to monitor key indicators of patient health (e.g. oxygen decompensation in Covid-19). The lack of clear guidance on remote monitoring was also identified.

At the patient level barriers include physical distance between patients' homes and the hospital, the stability of the patient's condition and their level of disability. Patients are selected using criteria which include their home environment, social support, health behaviour, and confidence in the care choice. High patient acceptance is a facilitator. Barriers included poor health literacy as well as internet access or affordability of technology Many of these factors will have implications for equity; equity-related concerns were the most common barriers in a review of remote monitoring technology in Covid-19. Models based on phone calls were considered more inclusive for patients without internet access or not comfortable with technology.

- **Q4; Q5 Models of care and use*:*** Hospital at home in studies included in evidence synthesis is used both as an alternative to hospital admission (step-up) and to shorten hospital admissions (step-down). In step-up use they may be used for people who attend the emergency department or those referred from primary or outpatient services. We did not identify reviews which were comparisons of different models of virtual wards, but the distinction between step-up and step-down models in evidence syntheses may not reflect clinical practice. While multidisciplinary teams are reported, most care is delivered by nurses and this is related to the level of non-acute care, as is family involvement.
  They are used predominantly for older people living in the community and people who have one or more chronic conditions, such as heart failure or COPD; people are treated both for exacerbations of those conditions and for unrelated acute illness such as stroke or pulmonary embolism. Many of these people would be considered frail. They are also used for people recovering after surgery and people at the end of life. This evidence is drawn from a range of systematic reviews, including Cochrane reviews of RCTs. However, trial populations may not reflect the population in clinical practice where services may be used for a wider group of people. People in the reviews were referred to step-up models from emergency departments, primary care, or outpatients. More recently virtual wards have been used for people with known or suspected Covid-19, but there is a lack of robust evidence in this population.
  A realist review identified four levels of factors related to successful models of remote monitoring:
    - Organisational (central dedicated units and workflow integration);
    - interpersonal (two-way communication, enhancement of self-management including medication adherence, personalisation and collaboration in care);
    - Intrapersonal (selection of patients with high risk of admission, motivation of both patients and staff, regular system-supported engagement to improve adherence);
    - Interventional (co-design with target population; making intervention simple/easy to use; accurate and sensitive measurements which are patient specific).

  A recent scoping review identified that patient-centred care is an aim of hospital at home but there is a lack of clarity about how this is planned and evaluated, and a lack of clear competence criteria for service delivery.

- **Q6. Experience:** There is low certainty evidence from Cochrane reviews that patient satisfaction may be improved by hospital at home in both step up and step down models. There is evidence from a well-conducted meta-synthesis of patient and other stakeholder experiences to support this: advantages identified included more comfortable and patient-centred care, more family engagement, and improved care. However, challenges identified included lack of round-the-clock patient supervision and increased caregiver burden. There may be increased caregiver stress and the need to involve and support caregivers was identified as important to prevent burnout. From a staff/service perspective the need for clear workflows with collaboration and coordination in care were identified as challenges. The need for institutional and organisational support for stakeholders was identified.

**Description of the intervention**

Virtual wards involve monitoring and treating people at home as an alternative to admission to hospital.(1) “Hospital at home” has been identified as a priority model for care.(2) People admitted to a virtual ward have the same access to (e.g.) diagnostic services as those admitted as inpatients. There are different models of virtual wards with varying levels of monitoring, visits and patient or carer involvement. We define virtual wards as any arrangement in which a person who would otherwise be admitted to hospital is managed at home with access to hospital services including monitoring. In this RES we consider virtual wards as an alternative to acute inpatient hospital admissions or stays and consider both step-up and step-down models. Virtual wards are included in 2018 NICE guidance on alternatives to hospital care.(3)

**Search**

We searched Medline, CINAHL and the Cochrane Database of Systematic Reviews. We also searched for preprints on MedRXiv. We also used citation scanning, particularly where we identified reviews of reviews. We used key terms “virtual wards” OR “remote monitoring” OR “hospital at home” for our main searches. Searches were carried out between 03 and 17 March; the main database searches were conducted on 16 and 17 March. An information specialist was consulted in determining the final approach to searching and carrying this out. We deduplicated and screened the searches from the main databases using Endnote.

**Key Questions**

We developed the following key questions with reference to key variables identified by the existing services in GM, and stakeholder consultation. For each of the key questions we looked in the first instance for existing evidence syntheses.

Q1. What is the effectiveness evidence for virtual ward admission compared to standard care (hospital inpatient care) for people who would ordinarily require acute hospital admission/stay? We looked to capture avoidance of acute inpatient hospital admissions and changes in length of inpatient stay in acute hospital wards (that is we considered virtual wards as either a substitute for (step-up model) or a complement to (step-down model) inpatient admission). We assessed effectiveness using the following outcomes: Length of stay in any setting (hospital or virtual ward), readmission, mobility, need for community support after discharge; achievement of rehab goals, adverse events including mortality, unplanned contacts/treatment events, acceptability (to patient/carer/staff), satisfaction (patient/carer/staff).

Q2. What is the cost-effectiveness evidence for virtual ward admission compared to standard care (hospital inpatient care) for people who would ordinarily require acute hospital admission? We considered all measures of cost-effectiveness (e.g. QALY) and relative cost-effectiveness (e.g. ICERs).

Q3. What are the barriers and facilitators for admission to a virtual ward? We considered factors at the patient and carer level and at the staff/system level. We noted where barriers include digital literacy of patients or carers. At the system level we also considered the barriers and facilitators of setting up virtual wards. We looked for evidence on the equity implications of identified barriers.

Q4. How are virtual wards being used? This is a broad question which addressed the following:

- What are the characteristics of people admitted and how long are they admitted for? This includes (e.g.) whether virtual wards are used instead of admission or as means of early discharge. We included equity considerations in these questions, looking at whether marginalised or vulnerable groups have a differential likelihood of being admitted to virtual wards. We accepted authors' definitions of marginalised or vulnerable but paid particular attention to BAME or low SES groups.

- What are the service contexts in which they are used?
- What level of patient engagement/input/activity (activation) is involved (what are the demands on the patient and how much feedback do they receive?)
- How intensive is the service provision and monitoring?

Q5. What is the evidence for the effectiveness and cost-effectiveness of different models of virtual wards compared to one another? We were specifically interested in evidence for the following:

- Different components and groups of components including but not limited to models with different levels of patient engagement (activation).
- Continuous vs intermittent monitoring. Models with different levels of patient engagement (activation)

Q6. What is the evidence for patient and carer experiences of admission to virtual wards? We accepted any measure of patient or carer experience, acceptability or satisfaction. We anticipated that this data might be qualitative or quantitative.

We used the following inclusion criteria for each question:

***Population:*** People who require acute inpatient hospital admission. We are particularly interested in the following prespecified groups of patients as well as those needing hospital admission more generally: people with a respiratory infection; people with heart failure; people with frailty. Where appropriate: carers for people who meet criteria. We included people who are admitted to virtual wards on discharge from inpatient care (as an alternative to a longer inpatient stay). We did not include people who are being treated as outpatients. People with long-term conditions were included where they would otherwise have been admitted to hospital as an acute inpatient.

***Intervention:*** Virtual wards (hospital at home). We define this as any arrangement in which a person who would otherwise be admitted to or remained in hospital is managed at home with access to hospital services including monitoring. We did not include studies of telemedicine services which are designed to provide virtual outpatient clinics or appointments as an alternative to in-person outpatient clinics, services or appointments; we also excluded remote monitoring of long-term conditions except where a hospital admission would be required (e.g. due to an exacerbation).

***Comparator:*** Standard care (admission to/remaining in hospital as an inpatient)

**Outcomes:** See individual key questions

***Study design:*** Evidence synthesis**.** We included all types of systematic reviews, including rapid reviews, and anticipated that the reviews we identified might not be limited to specific study types such as randomised controlled trials (RCTs); we included qualitative and mixed methods reviews as well as quantitative reviews. We did not include non-systematic narrative reviews of the literature. Reviews needed to have a documented search strategy and clear inclusion criteria to be included.

If we had not identified existing evidence synthesis for particular questions, outcomes or groups of interest, we would have looked at primary research and emphasised studies with the most robust research designs for the questions posed.

### RESULTS

#### Results of the search

Our search identified 630 records following database deduplication. We considered 52 of these at full text; we have drawn on 39 reviews identified through the search or reference checking. Because we identified a sufficient evidence base from existing evidence syntheses, we did not look for primary research studies. We are aware that there is overlap in the included studies across these reviews but have not used formal processes to account for this. Therefore, the evidence should not be treated as necessarily cumulative. This is a rapid evidence synthesis and therefore may not be comprehensive. Because evidence relating to interventions which were clearly virtual wards was limited we have drawn on the wider hospital at home literature.

#### Evidence for virtual wards

##### Q1: Effectiveness evidence for virtual wards

We identified several systematic reviews of virtual wards/hospital at home. Cochrane reviews were identified for the following indications:

- Hospital at home as step-up care(4, 5)
- Hospital at home as step-down care(6, 7)
- Hospital at home for COPD exacerbations(8, 9)
- Hospital at home for end-of-life care; may be relevant to some acute admissions.(10, 11)
- A general review of hospital at home (12-14) was superseded by the step-up and step-down care reviews.

Because Cochrane reviews are updated regularly, we identified multiple versions of some reviews; we cite the most recent. Cochrane reviews of interventions focus on effectiveness and safety but where relevant we have drawn on these for other questions, particularly those of cost-effectiveness and patient and carer experience. We have followed the Cochrane approach and reported results separately for the step-up and step-down models of care. These Cochrane reviews only include RCTs, and the more recent ones have a full GRADE assessment of the quality of the evidence, which we rely on here. Some of the hospital at home models may not be directly relevant to virtual wards as used in Covid-19 care.

###### *Step-up care*

Shepperd et al. 2016 looked at hospital at home for admission avoidance.(4) The authors included 16 RCTs (1814 participants). Included studies focused on people with COPD, people recovering from a stroke, mainly elderly people with acute medical conditions or various conditions. They found moderate certainty evidence that there was probably little difference in six month mortality (RR 0.77, 95% CI 0.60 to 0.99) or the likelihood of being transferred (or readmitted) to hospital (RR 0.98, 95% CI 0.77 to 1.23) while hospital at home may reduce the likelihood of living in residential care at six months' follow-up (RR 0.35, 95% CI 0.22 to 0.57). There was variation in the reduction of hospital length of stay.

We also identified a more recent review of hospital at home as an alternative to admission for patients with chronic disease, normally living in the community, who present at the emergency department.(15) The review included nine RCTs and found no difference between the groups in mortality (RR 0.84; 95% CI 0.61-1.15) or in functional status. Hospital at home patients were less likely to be readmitted to hospital (RR, 0.74; 95%CI, 0.57-0.95) and less likely to be admitted to long-term care (RR 0.16; 95%CI 0.03-0.74). Length of treatment was significantly longer in the hospital at home group (mean difference, 5.4 days; 95% CI 1.9-9.0 days).

A 2017 review looked at alternatives to hospitalisation in people aged over 65, and included 11 studies of hospital at home.(16) This looked at a range of conditions including COPD, heart failure, pulmonary embolism, pneumonia, stroke and uncomplicated diverticulitis as well as people living in the community or residential care with a variety of acute conditions. Most of the included studies were included in the Cochrane reviews identified here.(4, 8) However, the review included three RCTs in older people with a range of conditions including two where the home-care was delivered in the nursing homes where the participants lived.(16) All the trials were identified as being at high risk of bias and they were not pooled. Length of stay was shorter in the groups treated at home and other outcomes did not show significant differences.

A recent review of reviews evaluated effectiveness and safety of both step-up and step-down approaches. (17) This overview of reviews included ten reviews, nine of these were rated as high or moderate quality. In step-up models the reviews were reported to show a trend towards lower mortality (RR 0.77, 95% CI 0.54 to 1.09) and comparable or lower readmissions (RR 0.68 to 0.98).

Hospital at home was also included in a review of reviews of strategies to avoid inpatient hospitalisation for acute medical conditions.(18) This overview found that for several acute medical conditions, including heart failure and COPD, mortality rates and disease-specific outcomes were either improved or no different compared with inpatient admission. The reviews included in this section of the overview were identified and included here. An integrative review from 2014 looked at hospital at home in studies which recruited at least one third of their participants directly from the emergency department.(19) This included both randomised and non-randomised primary studies and earlier versions of the main Cochrane reviews as well as the Cochrane review on COPD.(8)

***Step-down care***

Gonçalves-Bradley et al. (2017) looked at hospital at home for early discharge.(7) They included 32 RCTs (N = 4746) which focused on people recovering from stroke, people undergoing elective surgery and people with a range of conditions. They reported these groups separately.

For people who are recovering from stroke they found there was probably little difference in three-to-six-month mortality (RR 0.92, 95% CI 0.57 to 1.48) and there may be little difference in the risk of hospital readmission (RR 1.09, 95% CI 0.71 to 1.66). Hospital at home may lower the risk of living in institutional setting at six months (RR 0.63, 96% CI 0.40 to 0.98). Hospital at home probably reduces hospital length of stay, the mean difference was seven days (95% CI 10.19 to 3.17 days).

For (mostly older) people with a mix of medical conditions there is probably little or no difference to mortality (RR 1.07, 95% CI 0.76 to 1.49) but hospital at home probably increases the risk of hospital readmission, although the results are compatible with both no difference and a relatively large increase in the risk of readmission (RR 1.25, 95% CI 0.98 to 1.58). Hospital at home may lower the risk of subsequently living in an institutional setting (RR 0.69, 0.48 to 0.99). There was probably a reduction in length of stay, but this ranged from 20 days to less than half a day.

In people with COPD there was insufficient information to determine the effect of hospital at home approaches on mortality (RR 0.53, 95% CI 0.25 to 1.12) but hospital at home may decrease the risk of readmission (RR 0.86, 95% CI 0.66 to 1.13); this was low certainty evidence in both cases. People experiencing an exacerbation of COPD were the focus of a Cochrane review from 2012; (8) as well as being included as a subgroup in this more recent review of step-down models.(7)

For people undergoing elective, mostly orthopaedic, surgery there may be no increase in mortality or readmission to hospital (low certainty evidence). Length of stay is probably on average four days shorter (mean difference 4.44 days, 95% CI 6.37 to 2.51 days earlier).

***Reviews including step-up and step-down care***

A recent review of reviews evaluated effectiveness and safety of both step-up and step-down approaches. (17) This overview of reviews included ten reviews, nine of these were rated as high or moderate quality. For step-down models the included reviews showed comparable mortality (RR between 0.92–1.03) and readmissions (RR between 1.09 to 1.25) to inpatient care and shorter hospital length of stay (MD between −6.76 to −4.44 days). A 2012 review of all types of “hospital in the home” included a very wide range of populations and interventions, including populations outside the scope of this rapid synthesis (e.g. psychiatric groups) and interventions which in many cases may not meet the criteria used here.(20) This found benefits in reduced mortality and readmissions across the groups and interventions assessed, but it is not clear how relevant these results are to this synthesis.

***Specific groups of patients: people with respiratory disease including COPD***

*People with known or suspected Covid-19*

We identified a rapid review and an integrative review focused on remote home monitoring for people with known or suspected Covid-19.(21, 22) Lack of comparison groups in the included studies is a limitation in both these reviews. A third review looked at technology for remote monitoring in people with Covid-19 but not virtual wards per se.(23)

The rapid review included 27 papers involving a wide range of models implemented in primary and secondary care sectors in eight countries for managing confirmed or suspected COVID-19 patients: 13 models functioned as pre-hospital admission, 5 as step-down wards, and 10 as both pre-hospital admission and step-down wards.(21) A range of methods were used to monitor people including telephone calls for paper-based reporting as well as more technological approaches such as mobile phone apps. Included studies were largely descriptive rather than comparative (no comparator was required in review eligibility criteria). The review reported on several relevant outcomes, but the data are of limited use because of the lack of a comparator. Mortality was reported as ranging from 0 to 3.1%. Escalation rates ranged from 5.1% to 26%; emergency department attendance/reattendance ranged from 2.5% to 36% and hospital admission/ readmission, ranged from 0 to 29%. Length of stay was reported as ranging from 3.5 days to 13 days; only one article reported reduction in length of stay, which was calculated at 5 days fewer per patient. The comparator for this was not reported.

The integrative review included nine case series studies and one retrospective nonrandomized controlled study were included; all 10 publications were rated as “good”.(22). Eight of ten studies gathered subjective (symptom reporting) and objective data (vital sign data) to remotely monitor patients; the most common data collected were oxygen saturation and temperature. All ten studies either used a smart phone application, a phone call, or a combination of both methods. Seven studies utilized an alert system that triggered a response from clinicians and the clinical team evaluated the need for escalation of care based on the participant’s data. This review did not summarise outcome data for key outcomes including ED admission and mortality. Patients were monitored for between 7 to 30 days, but it is not clear if these data represent length of stay.

The evidence for people with covid-19 is limited by the study designs in the identified reviews, which are not designed to answer comparative effectiveness or safety questions.(21, 22)

*People with COPD*

People experiencing an exacerbation of COPD were the focus of a Cochrane review from 2012; (8) as well as being included as a subgroup in the more recent review of step-down models (see above).(7) This review, which included all models of hospital at home, included eight RCTs (870 patients). The authors reported a reduction in readmission rates (RR 0.76; 95% CI 0.59 to 0.99) and a suggestion that there may be lower mortality (RR 0.65, 95% CI 0.40 to 1.04). For health-related quality of life the evidence was considered too weak to support firm conclusions. The evidence on mortality and readmission aligned with those from the COPD subgroup in the more recent review of step-down models of care with more certainty on readmission due to the larger number of trials/participants in the analysis.(7) Very similar results for mortality and readmission were also reported from a non-Cochrane review published in 2016. (24) The findings were also aligned with COPD evidence from a review of reviews where mortality RRs ranged from 0.65 to 0.68 and readmissions from RR 0.7 to 0.76).(17) An Ontario Health Technology Assessment(25) was carried out prior to the publication of the latest Cochrane review specific to COPD;(8) this drew on the earlier version,(26) and on earlier versions of the more general Cochrane reviews.

*People with newly diagnosed pulmonary embolism*

A 2012 review included eight studies including two RCTs in people with newly diagnosed pulmonary embolism without prior hospitalisation.(27) Ninety-day mortality was zero and from 741 people treated at home there were 13 recurrences and three major bleeding events, leading the review authors to conclude that this was a safe approach for selected patients. The single RCT which compared inpatient and outpatient treatment did not find differences between the groups but was underpowered to assess this as the events were uncommon.

*People with pneumonia*

A 2011 review compared outpatient with usual inpatient treatment of pneumonia and found comparable clinical outcomes and patient satisfaction, but it is not clear to what extent the outpatient treatment programs would meet the definition of hospital at home. Only one of the included studies was an RCT.(28)

***Specific groups of patients: people with heart failure***

We identified four reviews focusing on people with heart failure.(29-32) One of these looked at all models,(29) and another focused on post-discharge (step-down) models.(30) A review of step-up care for various conditions included only studies included in these reviews for heart failure.(16) A pair of reviews looked at transitional care arrangements to evaluate whether there is a dose-response relationship between intervention intensity and patient-centred(31) and healthcare-use;(32) these are indirectly relevant.

Qaddoura et al. (2015) included three RCTs and three observational studies.(29) Hospital at home did not alter all-cause mortality (RR 0.94; 95%CI 0.67 to 1.32) and showed a trend towards decreased readmissions (RR 0.68; 0.42 to 1.09); it increased the time to first readmission by two weeks (MD 14.1 days (95% CI 10.4 to 17.9)). These findings were supported by the observational studies. This is low to moderate certainty evidence based on the author's assessment of study quality.

Uminski et at. (2018) compared step-down models of hospital at home in people with heart failure to their use in people with undifferentiated high-risk chronic disease. (30) They found a reduced risk of both mortality (RR 0.59, 95% CI 0.44 to 0.78) and heart-failure-related readmission (RR 0.61, 95% CI = 0.49 to 0.76), (but not all-cause readmission) in people with heart failure; but they did not find these effects in people with high-risk chronic diseases generally.

The reviews of transitional care arrangements used meta-regression and dose-response analyses and found greater efficacy in trials that delivered the intervention by a multidisciplinary team while mortality and quality of life were improved and readmissions reduced with increased intensity and complexity of the transitional care interventions.(31, 32) This may be indirectly relevant to the review question.

***Specific groups of patients: people with frailty***

We did not identify any reviews which specifically focused on people identified as frail. However, many of the participants in the studies included in the identified reviews were elderly and/or had one or more chronic diseases. We identified a rapid review which looked at hospital at home as a component in the frailty pathway;(33) the evidence sources for hospital at home were overviews which included the Cochrane reviews of step-up and step-down care.(4, 7) This supports the suggestion that many of the people included in these reviews may be regarded as frail. The evidence from the RES as a whole is therefore probably relevant to people with frailty specifically.

A Cochrane review focusing on people at end-of-life deals with a subgroup of people who have a high level of frailty.(10) This review found high certainty evidence that home-based end-of-life care increased the likelihood of dying at home compared with usual care (RR 1.31, 95% CI 1.12 to 1.52); this is a more appropriate measure than mortality in this group. Admission to hospital varied among the trials (RR ranged from 0.62 to 2.61); other outcomes showed unclear effects.

We identified several reviews of transitional care arrangements, which can include hospital at home, which were focused on people who would be identified as frail. (34-36) These could be drawn on if indirect evidence were needed for this group.

**Q2: Cost-effectiveness evidence for virtual wards**

We identified a review of cost-analyses of hospital at home for acute conditions from 1996 to 2019, including 48 studies.(37) Most of the included studies found cost differences in favour of hospital-at-home but results varied from savings of €8773 to an increase of €2316 per patient. The authors assessed the studies using the Quality of Health Economic Studies (QHES) score. They found that the average score was 60 out of a maximum of 100 points and nearly all the studies failed to meet at least one of the criteria for risk of overestimation of cost savings. The most frequent problems were the use of average unit prices per inpatient day (not considering the decreasing intensity of care) and biased designs.

A recent review of reviews found that the cost implications of step-down models were unclear but there was a trend towards lower cost, compared to usual care, in step-up models.(17) This is in line with the Cochrane reviews: the review of step-up models found low certainty evidence that when the costs of informal care were excluded, admission avoidance hospital at home may be less expensive than admission to an acute hospital ward,(4) but the review of step-down models found it is uncertain whether hospital at home has an effect on cost in people recovering from stroke, or with a mix of medical conditions including COPD, or recovering from elective surgery (very low-certainty evidence).(7) Cost evidence was not always reported when sought – one review notes that no study reported out-of-pocket costs for patients or caregivers;(15) exclusion of these costs from evaluations may have implications for equity in the ability of people to access services.

For specific conditions, a rapid review in people with Covid-19 reported cost-related evidence such as cost per patient and the cost avoidances of using remote monitoring, this was non-comparative data and was not presented in detail; one study from England found that the mean cost per patient varied from £400 (step-down models) to £553 (step-up models).(21) The Cochrane review on people with COPD exacerbations assessed direct costs but considered that the quality of the available evidence was too weak to draw conclusions.(8) A non-Cochrane review from 2016 concluded that costs were lower compared to usual care in this population.(24) The Cochrane review of end-of-life care found that the effect on health service costs was uncertain (very low certainty evidence).(10) In people with heart failure one review found reduced costs in all three included RCTs.(29)

**Q3: Barriers and facilitators to virtual wards**

A recent review of reviews found that facilitators of implementation for virtual wards included coordinated and multidisciplinary hospital at home teams. Barriers to implementation included physical distance of patients' homes to the hospital, medical condition stability and level of disability. (17)

A recent meta-synthesis found that enablers for hospital-at-home development included clinicians with strong clinical and communication skills; supportive operational, regulatory and legal frameworks to promote care delivery in the home setting; and continuity of care including integration with post-discharge care.(38) Factors influencing patient selection of the hospital-at-home model included strong social support, positive health behaviours, confidence in receiving care at home, and conducive home environment. (38) These are all likely to have equity implications.

Similar barriers and facilitators were seen in reviews of virtual wards for Covid-19. In a rapid review, barriers and facilitators were not fully explored, but patient/carer training was identified as a determining factor of success.(21) No study targeted socially and economically disadvantaged groups, but models based on phone calls were considered more inclusive (i.e. including patients without internet access or technological literacy).(21) In a second review participant acceptance, feasibility, safety, and resource conservation were identified as key strengths while the barriers identified were more technical and organisational: lack of information on patient data security measures, robust outcomes testing, and identification of the most effective biomarkers to track oxygen decompensation.(22)

Many of these types of barriers, notably equity issues, lack of guidance and concerns around data protection and privacy were also identified by a rapid review of remote monitoring technology for people with Covid-19 which included 48 publications that described 35 distinct remote monitoring technologies; identifying perceived benefits of and barriers to their use.(23) Equity-related issues were the most commonly identified barriers. The review used mapping to PROGRESS-Plus to identify equity considerations: the most frequently reported characteristic was the "Plus" code for specific patient populations; followed by "place of residence", socioeconomic factors, race or ethnicity or culture or language, occupation, and gender or sex, education, and social capital; none reported on religion. Prominent themes reflecting barriers included equity-related barriers (e.g., affordability of technology for users, poor internet connectivity, poor health literacy). The review also identified the need for quality "best practice" guidelines for use of remote monitoring technology in clinical care, (this was the second largest group of barriers), and the need for additional resources to develop and support new technologies.

**Q4: How virtual wards are used**

**Q5: Different models of virtual wards**

We have combined these two questions as the evidence overlaps.

We identified two main models of virtual wards – use as step up models to avoid admission to acute care and as step down models to enable early discharge from acute care.(4, 7)

In the Cochrane review of step-up models the following definition was used:(4)

*"…hospital at home is a service that can avoid the need for hospital admission by providing active treatment by healthcare professionals in the patient's home for a condition that otherwise would require acute hospital inpatient care, and always for a limited time period. In particular, hospital at home has to offer a specific service to patients in their home requiring healthcare professionals to take an active part in the patients' care. If hospital at home were not available, then the patient would be admitted to an acute hospital ward."*

Patients were admitted from the emergency department, following primary care referral or by referral from outpatients. Care was provided by hospital outreach teams, a mix of outreach and community staff, or by general practitioners and community nursing staff. Care was provided by nurses, physiotherapists, social workers, with smaller numbers of studies reporting counsellors, cultural link workers and respiratory specialists, with pulmonary specialists available by telephone.

In the Cochrane review of step-down models the following definition was used: (7)

*"….hospital at home is a service that provides active treatment by healthcare professionals in the patient's home for a condition that otherwise would require acute hospital inpatient care, and always for a limited time period. In particular, hospital at home has to offer a specific service to patients in their home requiring healthcare professionals to take an active part in the patients' care. If hospital at home were not available then the patient would not be discharged early from hospital and would remain on an acute hospital ward."*

In these step-down models, intervention was delivered by hospital outreach services, community-based services, or was co-ordinated by a hospital-based stroke team or physician in conjunction with community-based services. The care provided was primarily nursing, with some additional care by care assistants or home helps. Specialist and dedicated nurses were involved as were occupational therapists, social workers, dieticians and speech therapists.

We identified a 2019 scoping review of patient-centredness, interprofessionalism and connectivity in models of care in the home including hospital at home.(39) While not a systematic review, this drew together and analysed relevant aspects of the services. The authors concluded that while patient-centred care is an aim of hospital at home there is a lack of clarity about how this is planned for and evaluated. They identified that the degree of non-acute care needs was inversely related to the level of interprofessional cooperation seen; higher levels of non-acute care were associated with more family involvement and co-ordination of care by nurses, with less involvement from other professionals. They identified a lack of clear competence criteria for delivery of home hospital services; elements reported included clinical experience and collaboration skills as well as formal qualifications. This evidence is limited by the fact that scoping reviews do not undertake quality assessment of the studies they include.

***Populations***

Some reviews only included studies which recruited patients who present to the ED and are older or have chronic disease.(15, 19) These also made up a substantial proportion of participants in wider reviews of step-up models.(4) We identified reviews which were specific to Covid-19, (21, 22) heart failure,(29, 30) COPD,(8, 24) stroke and people at the end of life, (10) while other reviews included studies which recruited people from these groups. A range of other acute conditions were represented in small numbers of studies. People who had undergone elective surgery were identified as a prespecified subgroup in a review of step-down models.(7) Most people included in reviews were older and living in the community. A review of activity levels in hospital at home found no eligible studies but did look at the activity levels of older hospital inpatients who would be eligible for hospital at home.(40) Sixteen included studies found that these patients spent 6.6% of their day active and had a mean daily step count of just 882.

The representativeness of trial (and hence review) populations in relation to those seen in routine clinical practice is not clear. As noted in barriers and facilitators (above) patient selection may be influenced by several actual or perceived barriers relating to patient and carer characteristics. In both clinical trials and practice these are likely to have equity implications and clinical trial inclusion criteria may be more restrictive than those used in service delivery.

In treatment of Covid-19 most models were for adult patients with COVID-19 symptoms (suspected and confirmed cases), including those over 65 years with significant comorbidities, or those with SpO2 above 92% at initial assessment. No study targeted socially and economically disadvantaged groups, but models based on phone calls were considered more inclusive (i.e. including patients without internet access or technological literacy).(21) These models were used primarily to identify early deterioration for patients self-managing COVID-19 symptoms at home (including those who had not been admitted to hospital and those who had been discharged), and to reduce the rate of hospital infection and demand for beds in the acute care sector. Teams managing patients in both step-up and step-down models varied in composition, but most were hospital-led with a minority either primary-care-led or managed jointly.

A paper by Jester et al. (2015) looked at models for evaluation of hospital at home services.(41) This was a review which also included a useful summary of the service being offered by Guy's and Thomas's in London; the authors' home institution. This outlined an approach which was flexible and operated as both a step-up and step-down service in practice; which may indicate that this is not necessarily a useful distinction in practice, outside the context of clinical trials. Given that the results of the effectiveness assessments in the two models are, in general, aligned, this may not be an issue in considering the generalisability of the findings.(4, 7)

***Factors involved in successful delivery***

We identified a realist review of factors influencing success of remote monitoring interventions.(42) This is indirectly relevant evidence because it relates to a broader class of interventions, but the factors associated with decreased and increased healthcare use (which was used as a measure of success) may also be relevant to virtual wards which incorporate remote monitoring as part of care. The authors identified four sets of factors associated with success in remote monitoring interventions: organisational; interpersonal; intrapersonal; and interventional.

Organisational factors were: central monitoring unit/dedicated professional for remote monitoring; integration of remote monitoring into workflow with a system to manage alerts; incentives to encourage uptake of remote monitoring.

Interpersonal factors were: encouragement of two-way interactive communication between patient and team; enhancement of self-management via support, education and feedback; use of data from the remote monitoring to personalise care; ensuring collaborative and multidisciplinary team involvement (including primary care).

Intrapersonal factors were: selection of patients at high risk of readmission (e.g. those with moderate to severe disease, high healthcare use, comorbidities); motivation of patients and staff to use remote monitoring; use of routine data entry checks and frequent follow-ups to increase adherence.

Interventional factors were: co-design with the target population; making the intervention simple and easy to use; ensuring accurate and sensitive measurements to enable early detection; using patient-specific measurements; enhancement of self-management (e.g. monitoring of medication adherence).

**Q6: Experience of virtual wards**

Chua 2022 qualitatively synthesised evidence on the perspectives of stakeholders who used the model of hospital-at-home.(38) This well-conducted meta-synthesis of 16 studies found that advantages of hospital-at-home include more comfortable and patient-centred care, more family engagement with patients, improved care continuity during and beyond hospital-at-home. From a service perspective the advantages are perceived better clinical outcomes for patients and increased hospital bed capacity. Challenges of hospital-at-home included lack of round-the-clock patient supervision compared to the hospital and increased caregiver burden. From a service perspective, disadvantages were unclear and underdeveloped workflows; difficulty in screening, identifying, and recruiting hospital-at-home patients; increased staff burden. This review concluded that high levels of satisfaction were expressed by various stakeholders. Continuity of care remains an important factor for patient-centeredness in hospital-at-home. Caregivers should be involved in the decision-making process and supported throughout the hospital-at-home duration to prevent caregiver burnout. Collaboration and coordination among healthcare professionals are vital and can be strengthened through training and technological advancements of remote patient monitoring. Institutional and organizational support for stakeholders may make hospital-at-home a viable solution to modern healthcare challenges. These themes were also seen in the scoping review by Vaartio-Rajalin (2019).(39)

Another recent review evaluated patient satisfaction.(15) Three studies that evaluated patient satisfaction reported mixed results: one study found a higher patient satisfaction in the hospital at home group than in the in-hospital group, whereas two studies showed no difference. This review also looked at caregiver stress, which may relate to caregiver acceptability. Two studies that evaluated caregiver stress reported mixed results: one found higher stress at admission that decreased at discharge in the hospital at home group, whereas caregiver stress did not change in the in-hospital group. The other study found no difference.

The Cochrane review of step-up hospital at home care found low certainty evidence that satisfaction with healthcare received may be improved with admission avoidance hospital at home but noted that few studies reported the effect on caregivers.(4) A review of reviews of interventions to avoid admission found patient and caregiver satisfaction with hospital at home that were either improved or no different compared with inpatient admission.(18) The Cochrane review of step-down hospital at home care found low certainty evidence that patient satisfaction may be slightly improved by hospital at home in people recovering from stroke, or elective surgery, or in those with a range of medical conditions.(7)

The Cochrane review of hospital at home for end of life care found low certainty evidence that it may slightly improve patient satisfaction at one-month follow-up, with little or no difference at six-month follow-up, but that the effect on caregivers and staff was uncertain (very low-certainty evidence).(10) In people with heart failure hospital at home improved health-related quality of life at both 6 months (SMD -0.31 95% CI -0.45 to -0.18) and 12 months (SMD -0.17 95% CI 0.31 to -0.02).(29) In people with Covid-19 a review found that only six of the models identified reported data on patient feedback, with high satisfaction rates (not reported in review).(21) A second review in this group concluded that the deployed remote patient monitoring programs were well accepted by participants.(22) In a review of remote monitoring technology, prominent themes associated with perceived benefits included a lower burden of care and reduced infection risk for hospitals and health care practitioners and provision of support for vulnerable populations.(23)

**Gill Norman with support from Chunhu Shi & Sophie Bishop (information specialist) April 2022**
